# Quantification of Bias-Variance Tradeoffs in Genome-Wide Polygenic Indices: When ‘Good’ PGIs are Bad and ‘Bad’ PGIs are Good

**DOI:** 10.64898/2026.11.18.26368405

**Authors:** Yavor Dragostinov, Rachel E. Hsu, Garrett W. Ennis, Diego Londono-Correa, Ted Schwaba, Michel G. Nivard, Elliot M. Tucker-Drob

## Abstract

Polygenic indices (PGIs) are downwardly biased in their predictive accuracies in independent samples due to the presence of estimation error in Genome-Wide Association Studies (GWAS) from which they are constructed. By performing GWAS of the same phenotype in independent samples, we obtain to error-corrected estimates of PGI prediction accuracy, which we use to estimate the extent to which different PGI construction methods (unadjusted, Clumping, LDpred2-inf, LDpred2-auto, SBayesR, PRS-CS-auto) index total single nucleotide polymorphism (SNP) heritability. To ensure practicality of this approach when individual-level data from the original GWAS are not available, we provide a means of synthetically splitting existing GWAS summary statistics into independent halves. In both simulated and empirical data, we find that advanced PGI construction methods that produce enhanced uncorrected prediction accuracies (often by shrinking individual SNP effects towards zero or exactly to zero) also produce deflated error-corrected prediction accuracies. LDpred2-inf, which most closely resembles a multiple regression estimator to account for linkage disequilibrium without further aggressive shrinkage, consistently produces PGIs with relatively lower uncorrected prediction accuracies but higher error-corrected prediction accuracies that nearly fully recover SNP heritability. We show that PGI construction methods that implement aggressive shrinkage may also provide biased patterns of associations with collateral phenotypes, irrespective of error correction. By providing a means of straightforwardly obtaining error-corrected estimates of prediction accuracy using PGIs derived from standard GWAS summary statistics, we have highlighted an uncommonly recognized bias-variance tradeoff associated with the choice of PGI construction methods.

## Introduction

A polygenic index (PGI; also commonly referred to as a *polygenic score* or a *polygenic risk score*) is a weighted combination of allele counts for many genetic variants into a single quantitative estimate of an individual’s genetic propensity for a trait or disease (Choi et al., 2020; Wray et al., 2007; Purcell et al., 2009). PGIs are most valid when applied to individuals from the population and environmental contexts matching the Genome-Wide Association Studies (GWAS) from which the weights used to construct them are estimated (Furrer & Turkheimer, 2026; Wang et al., 2026). However, even in such ideal situations, because of estimation error in the GWAS, PGIs are inherently noisy proxies for underlying genetic liability, and PGI prediction accuracy is substantially attenuated relative to the SNP heritability of the trait (Becker et al., 2021; Tucker-Drob, 2017; DiPrete et al., 2018). Further complicating PGI construction is that GWAS estimates represent marginal associations that do not account for linkage disequilibrium (LD) among variants. GWAS estimates are therefore ill-suited for direct use as PGI weights, for which conditional associations are more appropriate.

A variety of strategies have been developed to construct PGIs that both mitigate the effects of estimation error and account for LD among GWAS variants. The strategies differ from one another in their assumptions, mechanics, and degrees of sophistication, but can be generally classified as either sparsity-inducing (lasso-like; Tibshirani, 1996) or continuous (ridge-like; Hoerl & Kennard, 1970) shrinkage methods. Sparsity-inducing methods create PGI weights by shrinking GWAS estimates for a subset of variants to exactly zero, in effect creating a sparser set of genetic predictors. In addition to removing variants for which estimates are trivial relative to their uncertainty, sparsity-inducing methods remove highly correlated, effectively redundant, variants. Clumping and thresholding (e.g. PRSice-2; Choi & O’Reilly, 2019) is perhaps the most straightforward approach to inducing sparsity, whereas Bayesian mixture-based approaches (e.g. SBayesR; Lloyd-Pierce et al., 2019) that include a prior distribution composed of a point mass zero and one or more continuous distributions of effect sizes represent more sophisticated sparsity-inducing methods. Continuous shrinkage methods create PGI weights by shrinking GWAS estimates toward, but not exactly to, zero. One such continuous shrinkage method is PRS-CS which uses shrinkage to both adjust for LD and “adaptively squelch small noisy estimates towards zero, while leaving data-supported large signals unshrunk” (Ge et al., 2019, p. 2). In contrast, LDpred2-inf (Privé et al., 2020) assumes an infinitesimal model in which all variants have causal effects, and shrinkage largely functions to adjust for LD based on an LD matrix that is regularized to stabilize estimates.

Here, we identify an uncommonly recognized bias-variance tradeoff in PGI construction methods. Put simply, we find that methods that produce enhanced prediction accuracies (reduce error) also produce some of the most biased representations of genetic architecture. We reveal this phenomenon using a *split-half* method that divides the discovery GWAS sample into two independent halves in order to obtain and correct for measurement error in the PGIs, thereby producing an error-corrected estimate of PGI prediction accuracy. We also derive a pipeline for synthetically splitting existing GWAS summary statistics into two independent sets that both decreases computational burden and obviates the need for access to individual level data from the original GWAS or multiple independent GWAS of a trait of interest. Applications of these error correction approaches to both real and simulated data consistently demonstrates that the “best” PGI construction methods in terms of uncorrected prediction accuracy produce some of the most downwardly biased error-corrected estimates of prediction accuracy. We demonstrate that biased PGI construction methods may also evince biased patterns of associations with external phenotypes, even when implemented in standard settings (without error correction).

## Results

### Overview of Split Half Method for Error Correction

The split-half method for obtaining error-corrected estimates of polygenic prediction entails splitting the discovery GWAS sample into two approximately equally sized independent samples, conducting GWAS on each of these half samples, and using the resulting summary statistics to produce two PGIs (Tucker-Drob, 2017; DiPrete et al., 2018). Because the two PGIs are trained on the same phenotype but their estimation error is uncorrelated, their intercorrelation directly indexes the proportion of total variance in each that is true genetic signal (1 minus their intercorrelation indexes the proportion of total variance in each that is random measurement error). We use this property to estimate a simple structural equation model in an independent prediction sample that treats the two PGIs as indicators of an error-free latent variable, which is specified as a predictor of a measured phenotype. This model is represented general form by the path diagram in **Figure 1** and by the following system of equations

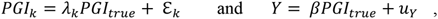

where *PGI*_1_ through *PGI_k_* are observed PGIs formed from k independent subsamples of the discovery GWAS sample, *PGI_true_* is an error-free latent variable, *Y* is the measured phenotype, ℇ*_k_* is the measurement error of *PGI_k_*, *β* is the structural regression of the *Y* on *PGI_true_*, and *u*_F_ is the residual of the regression of *Y* on *PGI_true_*. When the discovery sample is split into two equivalent independent halves, k takes on the values 1 and 2, and the constraint *λ*_1_ = *λ*_2_ is specified to represent the assumption that the two PGIs represent the latent variable equally well. We identify the scale of the latent variable in standardized units by specifying its mean to be 0 and variance to be 1. For ease of interpretation, *Y* and *PGI_k_* are standardized after residualizing for covariates (e.g. genetic principal components, age, and sex).

**Figure 1.**
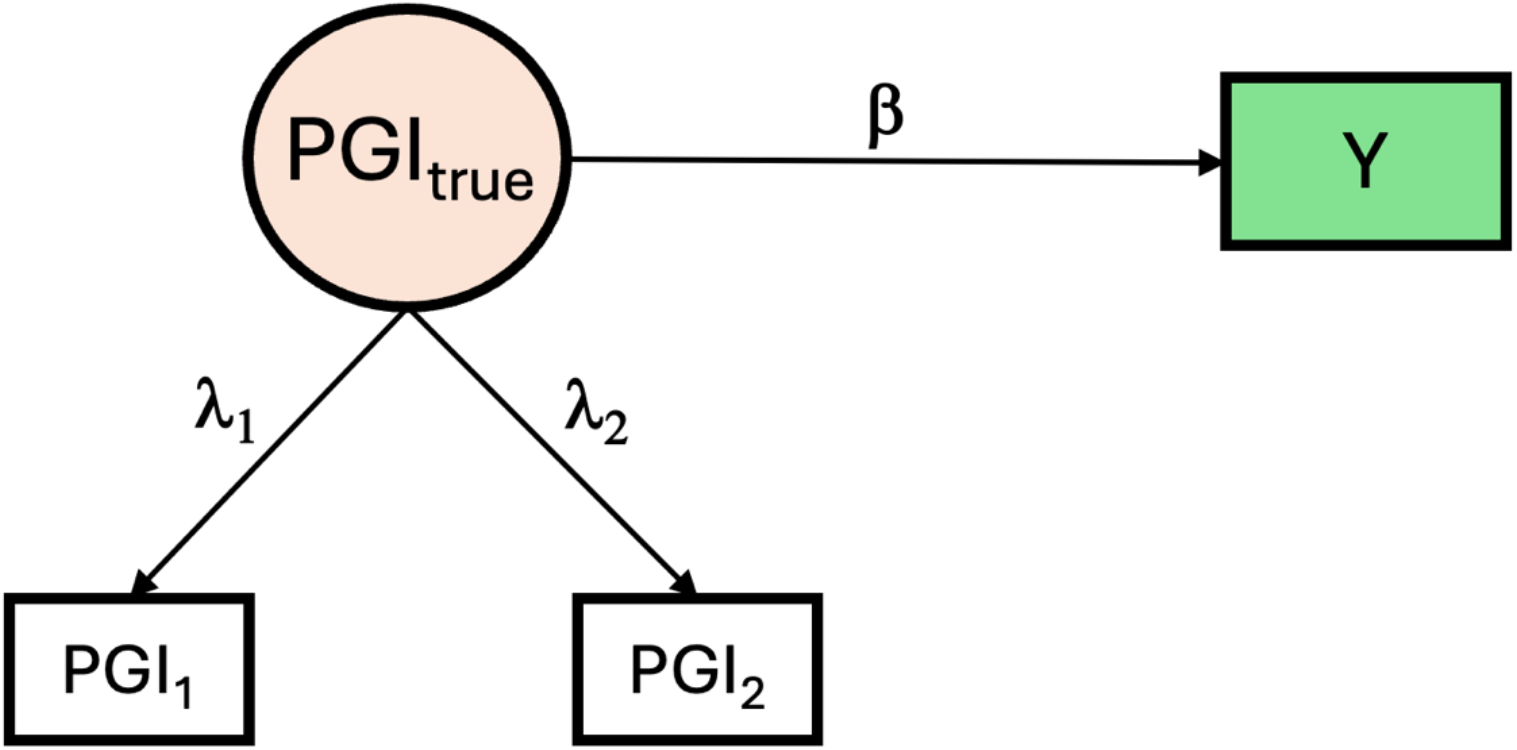
Path diagram of the structural equation model corresponding to the split-half model. *PGI*_1_ and *PGI*_2_ are two independent PGIs for the same trait constructed from independent, equally sized, subsets of the discovery GWAS sample. *PGI_true_* is a latent variable representing the true, error-free PGI. When the discovery sample is split into two equivalent independent halves, *k* takes on the values 1 and 2, and the constraint *λ*_1_ = *λ*_2_ is specified to represent the assumption that the two PGIs represent the latent variable equally well. The variance of PGI_true_ is fixed to 1 to identify its scale on the Z metric. The parameter *β* is estimated from structural regression of the *Y* on *PGI_true_*.

Under this model, the prediction accuracy *R^2^_PGItrue,Y_* from the regression of phenotype *Y* on the *PGI_true_* latent variable is an error-free index of the genetic variance in the phenotype indexed by the PGI. This term is a function of the squared observed correlation between each observed *PGI* and *Y (r^2^_PGIk,Y_)*, and the correlation between the two *PGI*s as follows:

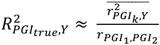

In practice, we estimate all parameters simultaneously in the system of equations constituting the structural equation model using maximum likelihood, from which we directly obtain our estimate of *R^2^_PGItrue,Y_* and its standard error using the delta method. Throughout this paper, we provide comparisons of the error corrected prediction accuracy (*R^2^_PGItrue,Y_*, which we also refer to as the error-corrected *R^2^*) and the mean of the uncorrected prediction accuracies of each half (*r^2^_PGIk,Y_*^-^). When PGI weights are unbiased, the prediction phenotype is the same as the discovery phenotype, and the prediction sample represents the same population (same genetic ancestry and distribution of age and environmental contexts) as the discovery sample, and the same set of SNPs that contribute to the SNP heritability of the trait (*h^2^*) are used to construct the PGI, the error-corrected *R^2^* is expected to be equal to SNP *h^2^*.

In many cases it may be difficult or impossible to access and process individual-level data from the original GWAS sample to compute GWAS in independent halves. We therefore also derived a pipeline for synthetically splitting existing GWAS summary statistics into two independent sets that both decreases computationally burden and obviates the need for access to individual-level data from the original GWAS sample (see **Methods**).

### Results of Simulations using Artificial Genotypes

As an initial test of the validity of the split-half method, we conducted a simple simulation with artificial biallelic variants in LD. By simulating using independent variants, this initial simulation focuses specifically on the problem of measurement error in the PGIs, and we construct PGIs directly from the marginal GWAS estimates. Our later simulations, which compare the performance of different PGI construction methods, are based on real genotypes, thus adding to the complexity associated with developing weights in the presence of both estimation error and LD. We drew *m* variants drawn from a two-trial binomial distribution with probabilities randomly drawn from minor allele frequency (MAF) = 0.05 to 0.50, resulting in *m* biallelic variants conforming to Hardy-Weinberg equilibrium with allele counts 0, 1, or 2 for *N* individuals. Standardized effect sizes for each variant on the phenotype were drawn from a normal distribution with mean 0 and variance = SNP *h^2^_true_/m*. We choose *m* = 60,000, corresponding to the effective number of independent markers in a standard GWAS (Wray et al., 2013), and selected SNP *h^2^_true_* = 0.20. Prediction sample size was fixed at 10,000 to produce stable estimates (prediction sample size affects uncertainty in estimates of prediction accuracy but does not affect the prediction accuracy expectation). We varied the discovery sample size from 20K to 3M (i.e., 10K to 1.5M per half) computing 5 replications per sample size condition. For comparison, we computed the average of the uncorrected *R*^2^ for the PGIs based on each half within each replication for each condition.

Results are shown in **Figure 2 and Supplementary Table S1**. We find that the error-corrected estimate closely recovers the SNP *h^2^_true_* across all discovery sample sizes, with trivial downward bias (mean range of *R^2^_PGItrue,Y_* estimates: 0.183–0.197; SNP *h^2^_true_* = 0.20) with standard errors decreasing as a function of discovery sample size (0.072 at *N* = 20K to 0.009 at *N =* 3M). In contrast, the uncorrected PGI accuracy follows the expectations of Daetwyler et al. (2008), asymptotically approaching SNP heritability with increasing *N*. We also confirmed that the same general pattern was obtained when non-effect variants were included in data generation to produce a spike and slab distribution. As expected, the error-corrected *R^2^* estimate remained unbiased under such a non-infinitesimal genetic architecture and uncorrected prediction accuracy was affected by the total number of independent variants (*m*) for which effects were estimated, rather than the total number of nonzero variants, with higher *m* associated with lower accuracy.

**Figure 2.**
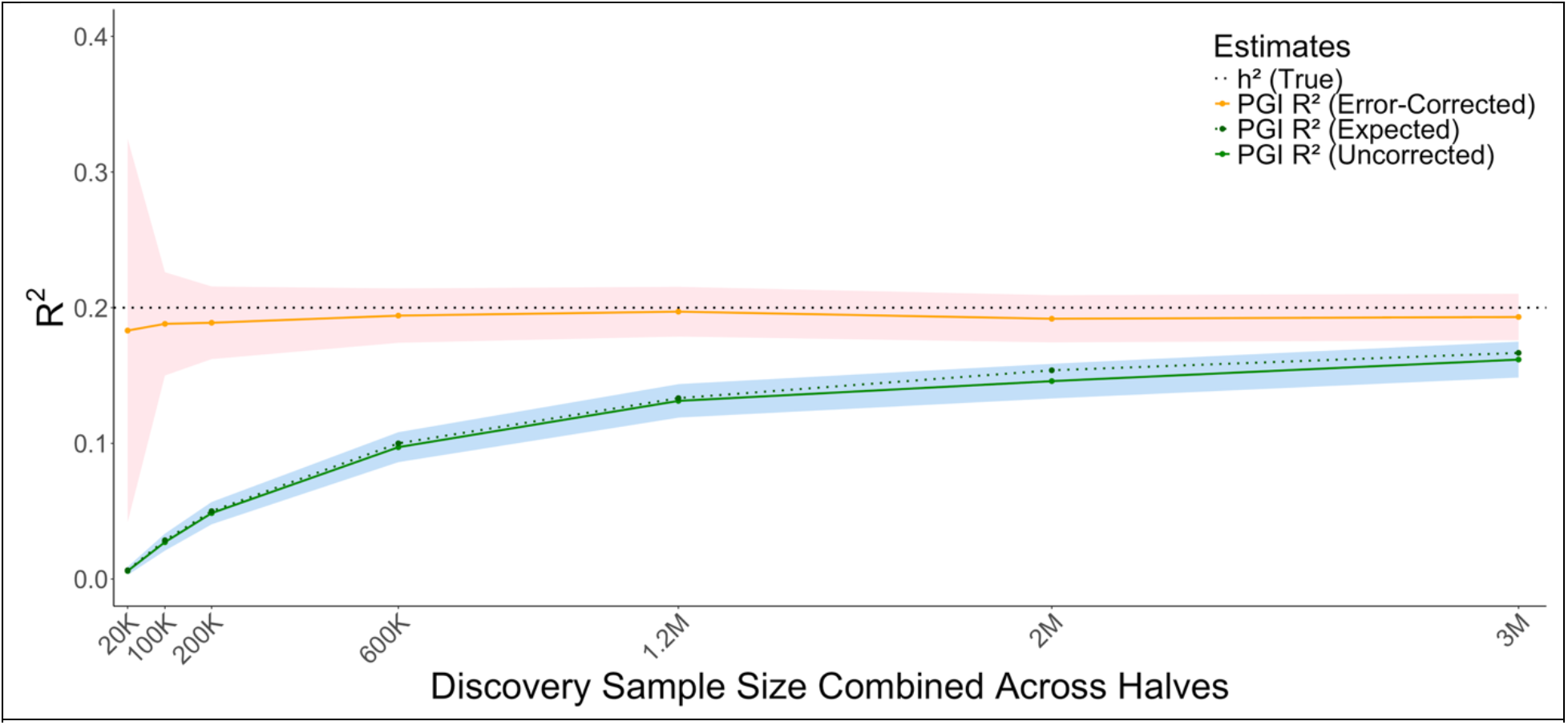
Simulation results. Observed and error-corrected PGI *R^2^* estimates at varying discovery sample sizes in simulations using artificial genotypes. The x-axis reflects the combined discovery sample size across both halves. The blue shaded band represents analytically derived 95% confidence intervals for the uncorrected *R^2^* estimates. The red shaded band represents the 95% confidence intervals for the corrected *R^2^* estimates. Corrected *R^2^* estimates closely match *h^2^_true_* at all sample sizes. Uncorrected PGI *R^2^* estimates track closely with theoretical expectations (Daetwyler et al., 2008), with prediction accuracy asymptotically approaching true heritability with increasing sample size.

### Results of Simulations using Real Genotypes

Having established that the error-corrected estimates closely recover SNP *h^2^* in the absence of LD among variants, we next conducted a series of simulations using real genotype data from the UK Biobank (UKB). Simulations were based on unrelated White British participants, and 749,619 HapMap3 variants present in UKB which passed standard genetic quality-control procedures, including filters on MAF, call rate, Hardy-Weinberg equilibrium, and imputation quality, and were also present in the LD reference panels used by the PGI construction methods. For each simulation replicate, a random subset of variants was designated as causal and used to generate a 100% heritable phenotype, to which normally distributed random variation then added to achieve the target level of heritability (e.g., 20%). Simulations were performed using genotypes from a discovery sample of 150,901 participants from UKB and genotypes from an independent prediction sample of 33,196 participants from UKB. The discovery sample was subsequently divided into two non-overlapping halves, and separate GWASs were conducted within each half to generate independent summary statistics for the split-half analyses.

#### Comparison of Six Popular PGI Construction Methods

We compared uncorrected *R^2^* and error-corrected *R^2^*estimates across PGIs constructed using unadjusted marginal GWAS estimates, Clumping, LDpred2-inf, LDpred2-auto, SBayesR, and PRS-CS-auto (see **Methods** section for details). We simulated the phenotype with SNP *h^2^_true_* = 20% and 20,000 causal SNPs. For each of the 5 replications, the discovery sample was divided into two non-overlapping halves and separate GWAS were conducted within each half, yielding ten GWAS in total. PGIs were subsequently constructed using each method, evaluated in an independent prediction sample, and the resulting estimates were averaged across replications. We do not explicitly consider the most recent methods that use functional annotations to further increase precision of estimates (e.g., SBayesRC, Zeng et al., 2024), or which combine GWAS estimates for a trait from multiple populations into a single PGI (e.g. PRS-CSx, Ruan et al., 2022). While such methods represent exciting progress in PGI construction, they retain the same core features of interest as are represented by the PGI construction methods considered here.

Results are displayed in **Figure 3** and **Supplementary Tables S2 and S3**. All PGI construction methods produced uncorrected *R^2^* estimates substantially below the SNP *h^2^_true_* = 20%; mean *R^2^_uncorrected_* = 3.8% to 7.5% across conditions). The error-corrected estimates were consistently higher, though several fell substantially below SNP *h^2^_true_*. LDpred2-inf produced the highest and most accurate corrected estimates (mean *R^2^_uncorrected_* = 0.188), followed by Clumping (SNP *h^2^* = 0.173), whereas PRS-CS-auto, SBayesR, and LDpred2-auto yielded more underestimated estimates ranging from 0.131 to 0.155, respectively. We observed a striking bias-variance-variance trade-off. The construction methods that produced the highest uncorrected *R^2^* estimates (PRS-CS-auto, LDPred2-auto, and SBayesR), appreciably exceeding the expectations of Daetwyler et al. (2008), produced the lowest (and most downwardly biased) corrected *R^2^* estimates. As expected, LD Score regression (LDSC) recovered true SNP heritability (SNP *h^2^_true_* = 0.20) precisely (LDSC *h^2^*= 0.200, SE = 0.008).

**Figure 3.**
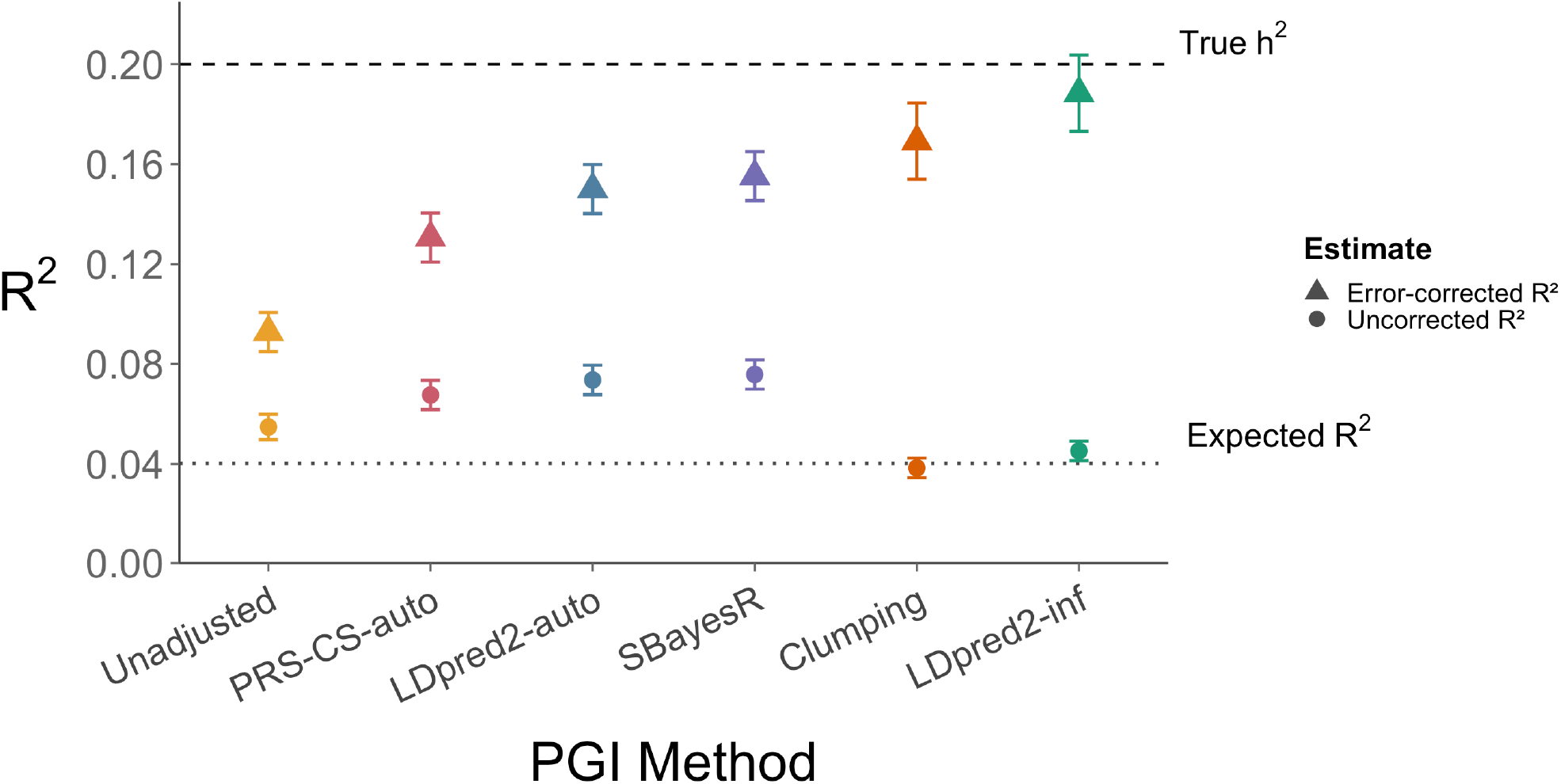
Simulation results. Comparison of uncorrected and error-corrected prediction accuracy across different PGI construction methods in simulations using real genotypes. Error-corrected estimates represent mean values across 5 replications per condition; error bars represent mean standard error per condition. The dashed horizontal line indicates the true simulated SNP heritability (SNP *h^2^_true_* = 0.20), and the dotted horizontal line indicates the expected predictive *R^2^* from the Daetwyler (2008) formula, assuming *M* = 60,000 effectively independent variants (Wray et al., 2013). Simulations were performed using genotypes from 150,901 participants from UKB for the discovery sample and genotypes from a separate set of 33,196 participants from UKB for the prediction sample.

#### Results Across Simulation Conditions Representing a wider range of Polygenicity and Genetic Architecture

We went on to examine the generalizability of the obtained pattern of results, focusing this more expansive set of conditions on SBayesR, LDpred2-inf, and Clumping. SBayesR was selected as an aggressive shrinkage-based estimator expected to yield the highest uncorrected *R^2^*but to be more biased after correction. LDpred2-inf was selected as an estimator that primarily serves to recover the multiple regression estimates without aggressively inducing shrinkage, and was expected to show lower uncorrected prediction accuracy but to be approximately unbiased and yield the most accurate corrected estimate. Clumping was included as a simple comparator.

We simulated under conditions in which we varied the true polygenicity from 500 to 100K causal SNPs (conditions = 500, 1K, 5K, 10K, 20K, 30K, 100K), and a condition in which 20K causal variants were sampled from the 21,190 SNPs in protein-coding regions using the baselineLD annotation model (Gazal et al., 2017). In these conditions, SNP *h^2^_true_* was set to 20%. In **Supplementary Tables S2** and **S3**, we also report a condition in which SNP heritability was set to 5% with and number of causal variants set to 20K.

**Figure 4** displays the distribution of estimates for each condition across the five replications. Several key features are evident. First, the advantage of SBayesR in terms of uncorrected prediction accuracy is accentuated at lower levels of polygenicity, with the uncorrected *R^2^* nearly matching the corrected *R^2^*at the lowest polygenicity condition (500) examined. By aggressively inducing sparsity in the PGI weights, SBayesR avoids including nonzero estimates for noncausal variants, which is especially advantageous when only a very small fraction of tested variants is causal. Nevertheless, across all conditions, SBayesR PGIs produce corrected *R^2^*estimates appreciably below both the SNP *h^2^_true_* and the corrected *R^2^* estimates from both clumping and LDpred2-inf. At increasing levels of polygenicity, LDpred2-inf continues to produce corrected *R^2^* estimates nearly matching the SNP *h^2^_true_*, whereas corrected *R^2^* estimates for both SBayesR and Clumping become increasingly downwardly biased. This can be understood because LDpred2-inf resembles a multiple regression estimator that includes all variants while taking into account their LD. In contrast, SBayesR aggressively induces sparsity, setting a potentially larger proportion of estimates for causal variants to 0 at higher levels of polygenicity. Similarly, clumping only allows for one nonzero variant per LD block, which is increasingly less likely to represent the true genetic architecture of the trait at higher levels of polygenicity.

**Figure 4.**
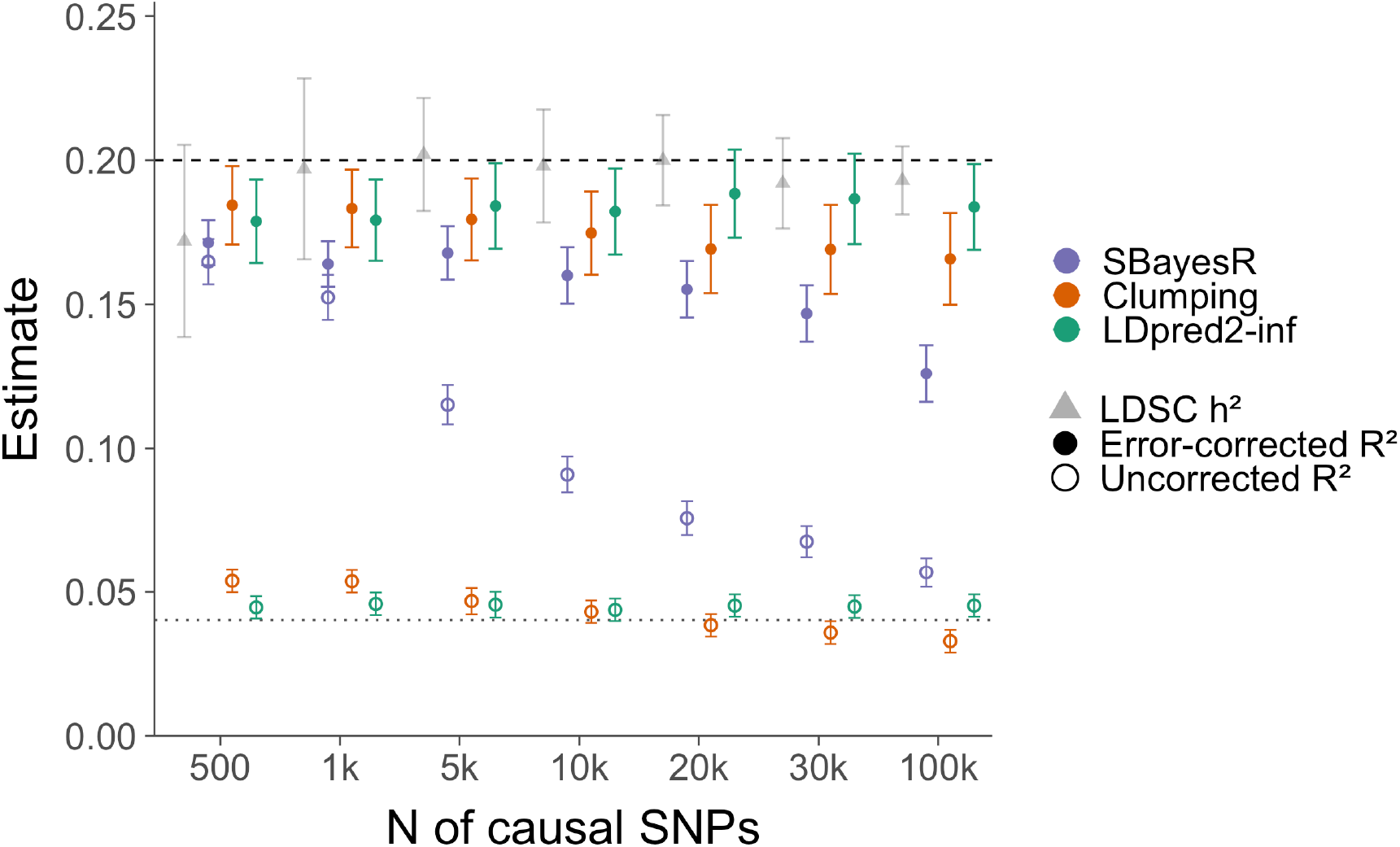
Simulation results. Error-corrected (filled circles) and uncorrected (open circles) PGI estimates by construction method, across simulated genetic architectures varying in polygenicity using real genotypes. Points and error bars represent means and mean SEs across five replications per condition. The triangles and error bars represent means and SEs of the LDSC *h^2^*across the five replications per condition. Dashed line = true simulated SNP *h^2^* (SNP *h^2^_true_* = 0.20). Dotted line = expected predictive *R^2^* from the Daetwyler (2008) formula, assuming 60,000 effectively independent variants (Wray et al., 2013). Simulations were performed using genotypes from 150,901 participants from UKB for the discovery sample and genotypes from a separate set of 33,196 participants from UKB for the prediction sample.

We additionally conducted simulations to evaluate the synthetic split-half approach focusing on LDpred2-inf. We examined conditions varying in polygenicity (10K, 20K, 30K causal SNPs) and in whether the noise GWAS and/or prediction sample were drawn from UKB or Health and Retirement Study (HRS). Error-corrected estimates from the synthetic split closely tracked the SNP *h^2^_true_* across all conditions, and were comparable to those obtained using real splits, while uncorrected *R^2^* remained noticeably attenuated regardless of configuration. Notably, using HRS as either the noise GWAS or prediction sample did not meaningfully influence the recovery of the SNP *h^2^_true_*, suggesting the synthetic approach is robust to cross-dataset application (see **Supplementary Tables S2** and **S3**).

### Results of Simulations of a Mixture Architecture using Real Genotypes

When a PGI is used to predict the same phenotype on which it was trained within an independent sample drawn from the same population, downward bias in its error-corrected *R²* relative to the total SNP *h^2^* of that phenotype may indicate that the PGI disproportionately represents different components of that trait’s genetic architecture. We simulated a scenario in which such bias might become apparent. Specifically, we generated a phenotype (AB) that was equally affected by a mixture of two distributions (50% of its heritability was attributable to each distribution) of causal effects, a relatively concentrated set of 10,000 causal variants (A) and a relatively diffuse set of 100,000 causal variants (B). Such a scenario broadly resembles that which might be expected for educational attainment (which has been modeled as arising from genetic components corresponding to cognitive and noncognitive factors; Demange et al., 2021) or molecular traits (e.g., testosterone, for which the genetic architecture is thought to reflect core genetic pathways superimposed on a far more highly polygenic background; Sinnot-Armstrong et al., 2021). We also generated phenotypes with genetic architectures arising from only one of these distributions each (A or B). We developed PGIs using both SBayesR or LDpred2-inf based on GWAS performed on the mixture phenotype (AB), which we then used to predict the non-mixture phenotypes A and B. The SNP *h^2^_true_* of each of the three phenotypes was set to 20%. If a PGI trained on the mixture phenotype (AB) is not disproportionately biased with respect to the two distributions of causal effects composing its genetic architecture, its associations with the non-mixture phenotypes (A and B) should be equal.

As hypothesized, we found that the SBayesR derived PGI for the mixture phenotype (AB) evinced a biased pattern of associations (both error-corrected and uncorrected) with the non-mixture (A and B) phenotypes (**Figure 5** and **Supplementary Table S5**). SBayesR derived PGIs were appreciably more strongly related to the phenotype created from the more concentrated polygenic architecture (A) than to the phenotype arising from the more diffuse polygenic architecture (B), despite both A and B phenotypes sharing exactly half of their respective genetic architecture with the mixture phenotype AB. In comparison, the LDpred2-inf derived PGI evinced an unbiased patterning of associations with the A and B phenotypes. Moreover, for LDpred2-inf, the error-corrected *R^2^* for both the concentrated phenotype A (0.093) and the diffuse phenotype B (0.093) each approached the theoretical maximum of 0.10. Of particular note is that the SBayesR model of genetic architecture produced biased patterns of association despite that it is explicitly a mixture model that subsumes the genetic architecture of the AB phenotype. In contrast, LDPred2-inf produced an unbiased pattern of associations despite the fact that its infinitesimal model of genetic architecture was a misspecification relative to the true model.

**Figure 5.**
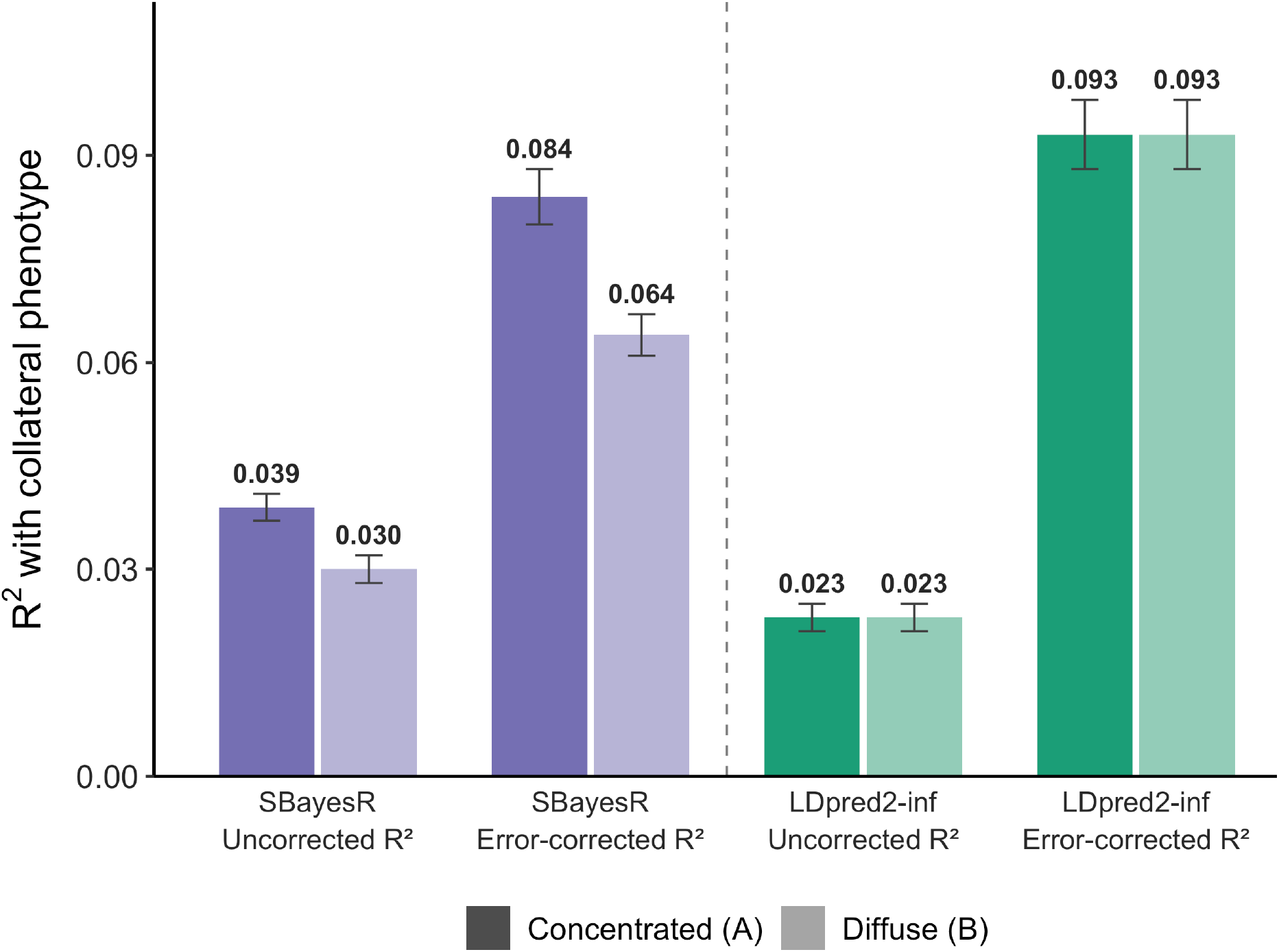
Simulation results. Error-corrected and uncorrected *R^2^* between the AB-derived PGI and collateral phenotypes A (concentrated architecture; 10K causal SNPs) and B (diffuse architecture; 100K causal SNPs), for LDpred2-inf and SBayesR in simulations using real genotypes. PGIs were constructed for the mixture phenotype AB using real UKB genotype data; A and B served as collateral outcomes with equal true genetic overlap with AB (theoretical maximum error-corrected *R^2^* = 0.10). Error bars represent SE of the mean *R^2^* across ten simulation replications.

### Results of Analyses of Empirical Data

Our primary empirical analyses were conducted using five phenotypes previously established to have highly polygenic architectures: educational attainment, height, body mass index, cognitive function (verbal-numerical reasoning), and neuroticism. We used independent subsets of the UKB dataset as GWAS discovery samples for training (which we in turn split into two independent half samples for split-half PGI construction) and prediction (**Table 1**). For neuroticism, we additionally used summary statistics from two independent approximately equally sized GWAS meta-analyses excluding UKB (Schwaba et al., 2026) to develop PGI weights, while maintaining the UKB prediction sample. PGIs were derived using SBayesR, Clumping, and LDpred2-inf. We compare uncorrected *R^2^* and error-corrected *R^2^* across methods, using LDSC-based estimates of SNP *h^2^* as benchmarks.

**Table 1.** Summary of Discovery GWAS.

| **Discovery Phenotype and Sample** | **Sample** | **N** | **M SNPs** | **Mean Chi Square** | **LDSC Intercept** | **Bivariate LDSC Intercept** |
|---|---|---|---|---|---|---|
| **Educational Attainment (EA)** | UKB (half 1) | 117,403 | 866,810 | 1.515 | 1.030 | 0.025 |
|  | UKB (half 2) | 117,393 | 866,810 | 1.531 | 1.032 |  |
| **Height** | UKB (half 1) | 118,671 | 866,810 | 2.370 | 1.119 | 0.098 |
|  | UKB (half 2) | 118,710 | 866,810 | 2.390 | 1.131 |  |
| **Verbal-Numerical Reasoning (VNR)** | UKB (half 1) | 49,596 | 866,810 | 1.254 | 1.027 | 0.009 |
|  | UKB (half 2) | 49,575 | 866,810 | 1.258 | 1.024 |  |
| **Body Mass Index (BMI)** | UKB (half 1) | 118,545 | 866,810 | 1.614 | 1.031 | 0.023 |
|  | UKB (half 2) | 118,564 | 866,810 | 1.613 | 1.021 |  |
| **Neuroticism (NEU)** | UKB (half 1) | 96,358 | 866,810 | 1.223 | 1.011 | -0.002 |
|  | UKB (half 2) | 96,318 | 866,810 | 1.243 | 1.013 |  |
| **Neuroticism (NEU)** | ReGPC excl. UKB (half 1) | 149,690 | 1,339,333 | 1.254 | 1.015 | -0.005 |
|  | ReGPC excl. UKB (half 2) | 186,107 | 1,348,559 | 1.371 | 1.007 |  |

Note. N = total number of participants. For ReGPC, N was constant across SNPs and the median N is reported. M = total number of SNPs. Mean Chi Square, LDSC Intercept, and the Cross-trait Intercept were estimated using LDSC from Genomic SEM using only HapMap3 SNPs.

Results are presented in **Figure 6** (see **Supplementary Table S6** for a detailed table with exact estimates and number of total and nonzero SNPs). Of the three PGI construction methods considered, SBayesR generally produced the highest uncorrected *R^2^* estimates and clumping generally produced the lowest. Error-corrected *R^2^* estimates were substantially higher than uncorrected *R^2^* estimates for the three PGI construction methods considered. Error-corrected *R^2^* from LDpred2-inf nearly fully recovered the SNP *h^2^* estimates from LDSC whereas error corrected *R^2^* from SBayesR fell substantially below the SNP *h^2^* estimates from LDSC.

**Figure 6.**
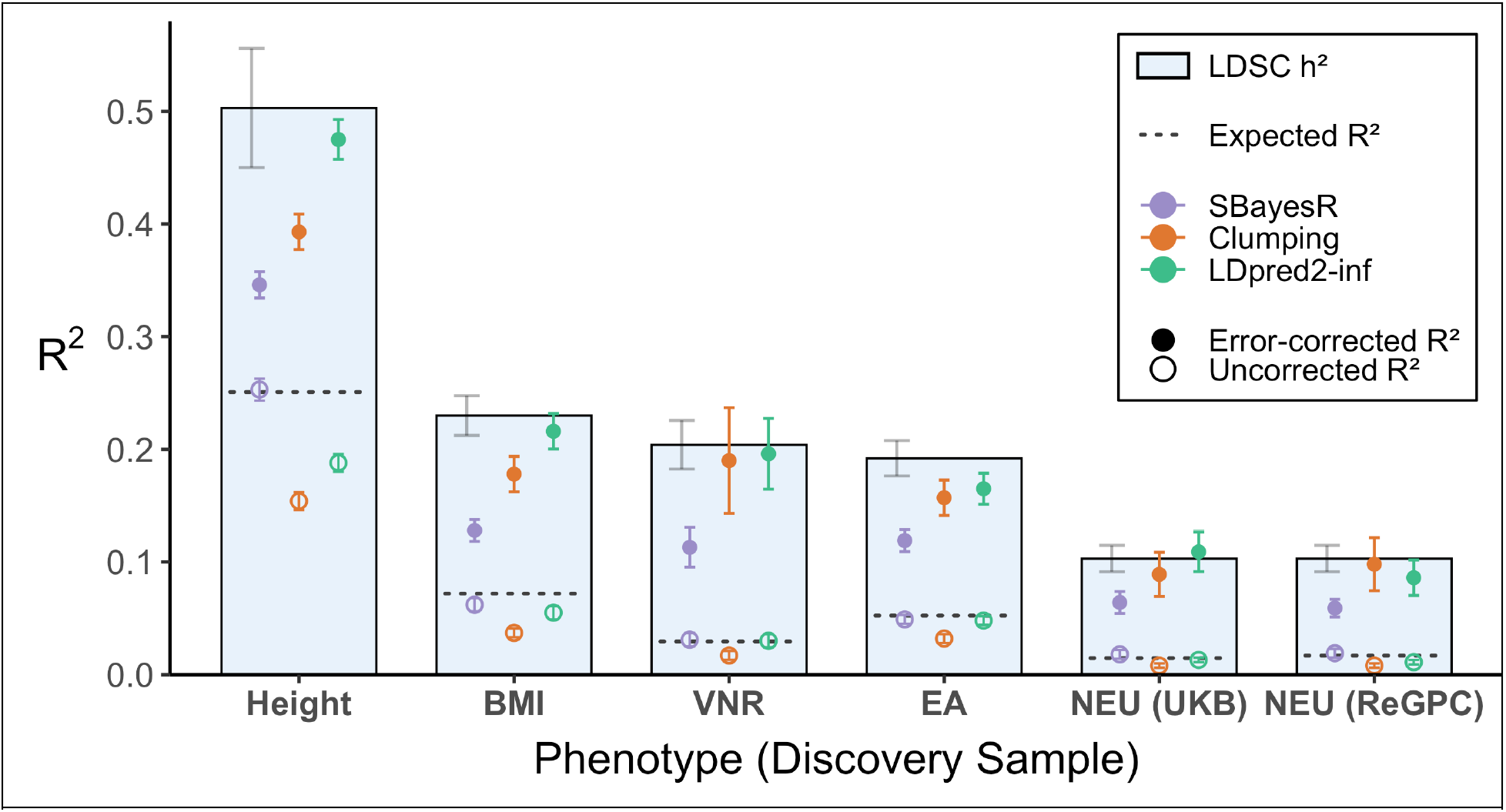
Empirical results. Error-corrected *R^2^* (filled circles) and uncorrected *R^2^*(open circles) for PGIs constructed using LDpred2-inf, Clumping, and SBayesR, inset within LDSC-based SNP *h^2^* estimates (bars), for six phenotypes. All PGIs were evaluated in a UKB prediction sample; NEU (ReGPC) differs from NEU (UKB) only in the discovery GWAS sample used to construct the PGI. Uncorrected *R^2^* values represent the average across the two half-specific models in which the observed phenotype was regressed directly on the corresponding PGI. LDSC SNP *h^2^* was estimated using GWAS summary statistics derived from the full UKB sample. Error bars on filled circles represent the SEs of the uncorrected *R^2^* estimates; error bars on bars represent the SEs of the LDSC SNP *h^2^* estimates. Dotted horizontal lines indicate the expected predictive *R^2^* from the Daetwyler et al. (2008) formula, assuming 60,000 effectively independent variants (Wray et al., 2013) and the corresponding discovery sample size for the corresponding PGI (Table 1); for NEU (ReGPC), the Daetwyler et al. (2008) formula was adjusted to allow discovery-sample SNP *h^2^* and prediction-sample SNP *h^2^* to differ.

We additionally applied the synthetic split procedure to existing summary statistics for educational attainment (Okbay et al., 2022, N = 689,700) and height (Wood et al., 2014; N = 253,280) from discovery GWAS that did not include UKB. We compared the resulting error-corrected *R²* estimates of LDpred2-inf PGIs to LDSC-based SNP *h^2^* estimates in the prediction sample, accounting for genetic correlations between the discovery and prediction phenotypes. The error-corrected estimates were broadly consistent with these theoretical benchmarks (**Supplementary Table S7).**

Having established that PGIs produced from LDpred2-inf provided corrected *R^2^* estimates most closely approaching SNP *h^2^*, we went on to compare results from applications using raw-data splits versus synthetic splits of the summary GWAS data used for training. LDSC-based estimates of genetic correlations for each pair of synthetic split halves were very close to 1.0 (mean *r*_g_ = 0.972), with cross-trait intercepts estimates very close to 0 (mean cross-trait intercept = 0.005), thus establishing the validity of the synthetic splits in producing independent GWAS summary statistics tapping the same true genetic signal. Results indicated that error-corrected *R^2^* estimates were highly similar across real and synthetic split halves (**Table 2**).

**Table 2.**
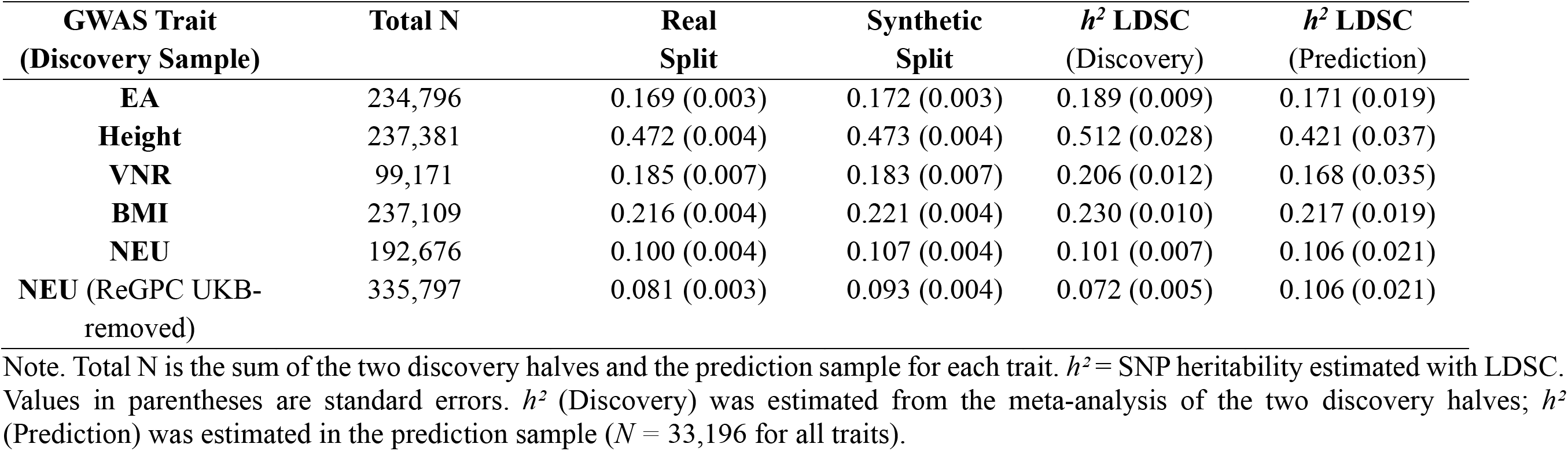
Error-corrected prediction accuracy estimates across corrected LDpred2-inf in real and synthetic split halves.

| **GWAS Trait<br>(Discovery Sample)** | **Total N** | **Real<br>Split** | **Synthetic<br>Split** | ***h*<sup>*2*</sup> LDSC**<br>(Discovery) | ***h*<sup>*2*</sup> LDSC**<br>(Prediction) |
|---|---|---|---|---|---|
| **EA** | 234,796 | 0.169 (0.003) | 0.172 (0.003) | 0.189 (0.009) | 0.171 (0.019) |
| **Height** | 237,381 | 0.472 (0.004) | 0.473 (0.004) | 0.512 (0.028) | 0.421 (0.037) |
| **VNR** | 99,171 | 0.185 (0.007) | 0.183 (0.007) | 0.206 (0.012) | 0.168 (0.035) |
| **BMI** | 237,109 | 0.216 (0.004) | 0.221 (0.004) | 0.230 (0.010) | 0.217 (0.019) |
| **NEU** | 192,676 | 0.100 (0.004) | 0.107 (0.004) | 0.101 (0.007) | 0.106 (0.021) |
| **NEU** (ReGPC UKB-removed) | 335,797 | 0.081 (0.003) | 0.093 (0.004) | 0.072 (0.005) | 0.106 (0.021) |

Note. Total N is the sum of the two discovery halves and the prediction sample for each trait. *h*<sup>*2*</sup> = SNP heritability estimated with LDSC. Values in parentheses are standard errors. *h*<sup>*2*</sup> (Discovery) was estimated from the meta-analysis of the two discovery halves; *h*<sup>*2*</sup> (Prediction) was estimated in the prediction sample (*N* = 33,196 for all traits).

## Discussion

In standard GWAS, associations for millions of SNPs with a phenotype are individually estimated with error. This presents two challenges for constructing PGIs. First, even small amounts of estimation error will compound across the myriad estimates, diluting or in some cases nearly completely overshadowing true signal. Second, the marginal associations obtained directly from GWAS do not account for LD among variants and must be weighted to approximate conditional estimates. PGI construction methods typically attempt to surmount both of these challenges. Methods that aggressively shrink individual SNP effects towards, or exactly to, zero can often increase prediction accuracy by reducing or removing SNPs whose true conditional effects are null or trivial relative to their estimation error. However, they may also produce biased representations of genetic signal. Using a combination of simulation studies and empirical applications, we find that after correcting for measurement error, many of the most predictive PGI estimation methods (LDpred2-auto, SBayesR, PRS-CS-auto) leave substantial portions of SNP *h^2^* unaccounted for. We find that LDpred2-inf, which most closely resembles a multiple regression estimator to account for LD, produces PGIs with lower realized prediction accuracy but nearly fully recovers SNP *h^2^* after error correction.

By providing a means of obtaining error-corrected estimates of prediction accuracy, we have highlighted what we believe to be an uncommonly recognized bias-variance tradeoff in the derivation of PGIs. Researchers are likely widely aware of this trade-off when implementing clumping and p-values thresholding methods that explicitly remove all but one SNP within a locus and all SNPs falling above a particular p-value significance threshold. For instance, it is well-known that when constructing PGIs exclusively from independent genome-wide significant loci, the resulting index will only represent a portion of genetic propensity for polygenic traits and that weakly associated SNPs may exhibit different patterns of association with collateral traits than is exhibited by the PGI. Such limitations may be more difficult to intuit when using an advanced method that relies, for example on a mixture distribution. Nevertheless, a straightforward way to understand the results is as follows: construction methods that rely on aggressive shrinkage to obtain enhanced prediction accuracies, do so by discounting or eliminating the contributions from some components of polygenic architecture, thereby providing biased indices of polygenetic effects on the phenotype that the PGI was trained to index.

Our simulation results highlight that the same general pattern of bias-variance tradeoff applies across a wide range of trait polygenicities. One might expect bias to occur when sparsity-inducing PGI construction methods are applied to traits with a near infinitesimal polygenic architecture. However, we find that mixture-model based PGI construction methods suffer from the same limitations even when a relatively smaller number of causal variants contribute to polygenic signal. In fact, even though the mixture model assumed by the SBayesR model subsumes the model used to generate the data in our simulations (i.e., we generated SNP effects from a mixture of a normal distribution of causal effects and a point mass of noncausal effects), PGIs produced using “incorrect” LDpred2-inf model were produced error-corrected estimates of prediction accuracy that were higher, and closer to total SNP *h^2^*. We attribute this to the inability of the mixture model to perfectly determine the identity of causal variants, thereby setting estimates for some causal variants to 0 rather than including their noisy signal in the PGI. In contrast, LDpred2-inf operates primarily a multiple regression estimator that includes all variants while accounting for their LD, with regularization performed on the LD matrix to stabilize estimates without otherwise aggressively shrinking estimates. While LDpred2-inf bears its name to reflect its characterization as an infinitesimal model in which all variants are assumed to have a nonzero effect, it might be best to view it less as an infinitesimal model, but as a model that allows each variant to have a nonzero estimate without imposing aggressive shrinkage. Importantly, our findings illustrate that the optimal PGI construction method depends on its intended application. When the primary objective is to maximize prediction accuracy, methods that introduce systematic bias may nevertheless be preferable because of their lower variance. However, when the objective is to capture the full genetic architecture of a trait, minimizing systematic bias may be more important than maximizing prediction accuracy.

Our correction of prediction accuracy for measurement error in the PGIs builds directly on work by Tucker-Drob (2017) and DiPrete et al. (2018) and on classic methods from psychometrics and econometrics (Spearman, 1904). In recent years, genetic research implementing various approaches to error-correction methods has become increasingly common (Becker et al., 2021; Frach et al., 2026; Malawsky et al., 2026; Pingault et al., 2021). However, previous applications have not considered the important role that the choice of PGI construction method plays. In fact, rather than treating the error-corrected estimate of prediction accuracy as a quantity to be estimated independently of SNP *h^2^* (as we have done here), some methods (e.g. Becker et al., 2021, Pingault et al., 2021, Frach et al., 2026) use an external estimate of SNP *h^2^*as a fixed value to identify the error correction model. Our results indicate that this choice may bias downstream estimates, especially when using aggressive shrinkage-based PGI construction methods. An advantage of methods that use an external of SNP *h^2^* has been that they can implement existing GWAS summary statistics and do not require access to individual-level data from the GWAS sample. Such data are often difficult or impossible to access, such that directly splitting the discovery sample can be impractical. Here, we have derived method for synthetically splitting GWAS summary statistics to provide a means of obtaining error-corrected estimates of prediction accuracy without relying on the strong assumption that the external estimate of SNP *h^2^* generalizes to the PGI method(s) employed or to the prediction sample. We expect that there may be wide-spread applications of the synthetic split approach that we have introduced here beyond the estimation of obtaining error-corrected estimates of prediction accuracy. To facilitate implementation of synthetic split approach, we are making publicly available noise GWAS summary statistics generated using genotypes from the UKB (*n* = 49,879) and the HRS (*n* = 9,878).

SNP *h^2^* estimates are often interpreted as representing the theoretical upper asymptote of PGI prediction with increasing GWAS discovery sample sizes (Wang et al., 2023). Error-corrected estimates of PGI prediction cannot be universally interpreted in this way. For many PGI construction methods, the extent of shrinkage may vary systematically with GWAS discovery sample sizes, such that PGIs derived using larger GWAS exceed the error-corrected prediction accuracies of PGIs derived for the same trait using smaller sample sizes. For many PGIs for which error-corrected prediction accuracies appreciably differ when implementing moderately-size GWAS discovery samples, prediction may converge with one another and with SNP *h^2^* as GWAS sample sizes approach levels needed for saturation. Of course, PGI prediction may also exceed SNP *h^2^* for other reasons, such as the inclusion of rare variants in the PGI that are not included in the estimate of SNP *h^2^*, estimation of SNP *h^2^* using GWAS meta-analysis of heterogeneous samples, and employment of a more reliable measure of the phenotype in the prediction sample than was used in the discovery sample.

In summary, we have used both simulation and empirical application to obtain error-corrected estimates of prediction accuracy of PGIs constructed using a variety of construction methods. Our results highlight an uncommonly recognized bias-variance tradeoff in the choice of PGIs. PGIs produced by methods that aggressively shrink information from a subset of variants (LDpred2-auto, SBayesR, and PRS-CS-auto) may not represent the broader genetic architecture of the phenotype and may evince biased patterns of associations with collateral phenotypes. LDpred2-inf, which most closely resembles a multiple regression estimator to account for LD, produces PGIs with relatively lower prediction accuracy but after error correction nearly fully recovers SNP *h^2^* and produces unbiased patterns of associations with collateral phenotypes. Importantly, while we have used error correction to index and highlight this phenomenon, this limitation applies to results from standard applications of PGIs within statistical genetics, even when error correction is not of interest. Nevertheless, when the primary goal is to maximize individual-level prediction, aggressive shrinkage-based methods may still be the most appropriate choice.

## Methods

### Protocol for Synthetically Splitting GWAS Summary Statistics

In many cases it may be difficult or impossible to access and process individual-level data from the original GWAS in order to compute GWAS into independent halves. We derived a pipeline for synthetically splitting existing GWAS summary statistics into two independent sets that both decreases computationally burden and obviates the need for access to individual-level data from the original GWAS (see **Extended Data Figure 1** for a conceptual overview of the protocol). Here, the noise GWAS refers to a GWAS of a pseudo-phenotype randomly sampled independently for each individual from a standard normal distribution (*M =* 0, *SD =* 1). The empirical GWAS is likewise conducted on a standardized phenotype (*M =* 0, *SD =* 1), so that the scale of the empirical phenotype matches that of the randomly generated pseudo-phenotype. The noise GWAS sample may overlap with the discovery GWAS sample without consequence, as the noise phenotype is, by construction, uncorrelated with the true phenotype. Both the true and pseudo-phenotype GWASs must be conducted using genotypes from individuals that are not included in the prediction sample.

The sample size of the pseudo-GWAS does not need to correspond to that of the empirical GWAS. In order to place the noise effect estimates on the appropriate scale, a per-SNP scaling coefficient is computed as:

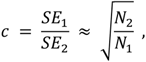

where SE_1_ and SE_2_ are the per-SNP standard errors from the empirical and noise GWAS, respectively, and N1 and N2 are their corresponding sample sizes. The two synthetic halves are then constructed as:

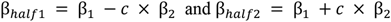

By construction, β_half1_ and β_half2_ share the same expected genetic signal but are affected by independent sampling variation, thereby approximating the statistical properties of two independent GWAS conducted on the same phenotype. While it would be straightforward to randomly generate noise to directly add and subtract from GWAS beta, performing a pseudo-GWAS produces disturbances that are appropriately correlated across SNPs as a function of LD, which is required to obtain an appropriate synthetic split.

### Software

GWAS for the simulation analyses were conducted in PLINK2 (Chang et al., 2015), whereas all empirical analyses GWAS were conducted using REGENIE (Mbatchou et al., 2021) to account for the genetically related individuals in UKB. Phenotypes for the simulations were generated using GCTA (Yang et al., 2011). Subsequent statistical analyses were conducted in R, including estimation of LDSC SNP *h^2^* using Genomic SEM (Grotzinger et al., 2019), estimation of PGI weights using LDpred2-inf implemented in the bigsnpr package (Privé et al., 2020), and structural equation modelling analyses were conducted using the lavaan package (Rosseel, 2012). In addition, SBayesR (Lloyd-Jones et al., 2019), implemented in the GCTB software (Zeng et al., 2018), and clumping were done in PLINK2; all PGIs were scored using PLINK2. Computationally intensive analyses were run on the Texas Advanced Computing Center (TACC) Lonestar6 cluster, with pipelines managed using Bash.

### Simulations

We conducted a series of simulation analyses using individual-level genotype data from the UKB and the HRS to evaluate the performance of PGI construction methods. Across all conditions, we simulated quantitative phenotypes under a genetic architecture with varying numbers of causal SNPs (500, 1K, 5K, 10K, 20K, 30K, 100K, and a 20K condition restricted to functional annotations) and two levels of SNP heritability (SNP *h^2^_true_* = 0.20 and SNP *h^2^_true_* = 0.05).

For each condition, we ran five independent replications, resulting in a total of 120 simulations. In each replication, causal SNPs were randomly sampled from the available genotype data, and their effect sizes were generated using GCTA. Individual-level genetic values were computed as the sum of allele dosages weighted by these true effects. Phenotypes were constructed by adding normally distributed environmental noise such that the target heritability was achieved. All phenotypes were subsequently standardized.

For the functional annotation condition, causal variants were restricted to protein-coding regions using the baselineLD annotation model (Gazal et al., 2017). Specifically, we first selected SNPs annotated as coding in the baselineLD files (approximately 39,000 SNPs), then intersected this set with SNPs present in our genotype data (resulting in a total of 21,190 SNPs). From this subset, 20,000 SNPs were randomly sampled as causal variants in each replication.

#### UKB GWAS

Genome-wide association studies (GWAS) were conducted in UKB using a total of 150,901 White British unrelated participants using the simulated phenotypes. Prior to analyses we implemented quality-control (QC) specifications such as excluding SNPs with Minor Allele Frequency (MAF) < 1%, call rate < 95%, and those that deviated from Hardy-Weinberg Equilibrium (*p* < 1×10^-6^). SNPs with poor imputation quality (INFO < .40) were removed, while all SNPs were restricted to HapMap3. The sample was split into two non-overlapping halves, and GWAS were performed separately within each half while controlling for covariates (sex, age, age squared, birth cohort, and genetic principal components). This produced two independent sets of summary statistics per replication.

#### Construction of Synthetic Discovery Samples

In the real split condition, PGIs were derived directly from the two independent GWAS halves. In the synthetic split condition, summary statistics from the two halves were meta-analyzed to form a combined discovery sample, which was then artificially split into two independent halves. PGI weights were derived from each half using multiple methods, including LDpred2-inf, SBayesR, and Clumping.

#### UKB Prediction Sample

We selected a random subset of 30,000 White British participants and ensured they did not overlap with the UKB discovery samples, nor include related individuals (kinship > 0.05) across either half of the prediction sample.

#### HRS Phenotype Construction and Prediction

To assess predictive performance, matched phenotypes were constructed in HRS using the same causal SNPs and effect sizes as those used in the UKB simulations. Specifically, the true effect sizes from the UKB simulation step were mapped to HRS SNP identifiers and used to compute individual-level genetic values in HRS. Environmental noise was then added to achieve the target SNP heritability (SNP *h^2^_true_* = 0.20) and phenotypes were standardized.

PGIs derived from UKB GWAS (both real and synthetic splits) were subsequently applied to the HRS genotype data (*n* = 9,878 EUR participants) to generate predicted scores. Because HRS phenotypes were constructed using the same underlying genetic architecture as the UKB simulations, this framework enables direct evaluation of predictive accuracy and recovery of the true genetic signal under known conditions.

### Empirical Analyses

#### Population-level GWAS

The primary GWAS discovery sample was a subset of White British participants from the UK Biobank (*n =* 237,944).

#### UKB Prediction Sample

Our primary prediction sample was a subset of white British UKB participants not included in the discovery sample (*n* = 33,195). To prevent sample overlap and cryptic relatedness, we excluded any prediction-sample participant with a kinship coefficient > 0.05 with any participant in either discovery half, which corresponds roughly to third-degree relatives or closer. Kinship was estimated with the UKB relatedness file (Bycroft et al., 2018).

#### UKB Discovery Sample

The primary GWAS discovery sample was a subset of White British participants from the UK Biobank (*n* = 237,944) who were not included in the prediction subsample. Genetically related individuals were identified using UK Biobank’s pre-computed pairwise kinship coefficients. Participants were clustered into family units by grouping all individuals connected through kinship relationships (kinship > 0.05), such that individuals related either directly or indirectly (via intermediate relatives) were assigned to the same family cluster. Individuals with no detected relatives were treated as single-member families and assigned unique family identifiers. The same kinship information was additionally used to ensure that no individual included in the discovery sample was related (kinship > 0.05) to any individual in the prediction sample, thereby preventing cross-sample genetic relatedness.

To construct independent split-half discovery samples while preserving family structure, the split was performed at the family level rather than the individual level. First, one row per family was created and the number of individuals in each family was recorded. The order of families was then randomly shuffled to prevent any systematic differences (e.g., due to recruitment order or family size) from influencing the split. Following this randomization step, families were sequentially assigned to the first half in their shuffled order until the cumulative number of individuals in half 1 reached approximately 50% of the total sample size. All remaining families were assigned to half 2. This ensured that no related individuals were split across halves, and the two halves were approximately equal in sample size, while the initial random shuffling ensured that assignment to half 1 versus half 2 was not systematically structured.

GWAS were then conducted separately for each phenotype in each split-half sample using REGENIE, with sample size ranging between 49,596 and 118,671 across halves (see **Table 1**), respectively. LDSC confirmed near-perfect genetic correlations across the two split-half GWAS for each (mean *r*_g_ = 1.014), and independence of estimates (mean cross-trait intercept = 0.012).

#### UKB GWAS Phenotypes

Within UKB, we curated educational attainment (EA; UKB data-fields 6138 and 845), standing height (data-field 50), body mass index (BMI; UKB data-field 21001), verbal numerical reasoning (VNR; labelled as “fluid intelligence score” in the UKB; UKB data-field 20016), and neuroticism (UKB data-field 20127) in UKB. For each phenotype, the discovery sample was split into two halves, and separate GWAS were conducted in each half.

#### UKB GWAS

The same QC procedures described for the discovery sample in the simulations were implemented prior to association testing. For each trait of interest, we regressed the phenotype on each available autosomal biallelic SNP, controlling for chromosomal sex, age, age^2^, birth cohort, and 10 genetic principal components (PCs) for ancestry. All phenotypes were standardized prior to association testing.

#### External GWAS Summary Statistics

We used two independent approximately equally sized meta-analyses of neuroticism from the Revived Genomics of Personality Consortium, with UKB left out (ReGPC; Schwaba et al., 2026), which we treated as split-half GWAS summary statistics for error correction analyses (*N* = 149,690 for half 1, *N* = 186,107 for half 2). Additionally, we applied the synthetic split procedure to GWAS summary statistics for educational attainment (Okbay et al., 2022, *N* = 689,700) and height (Wood et al., 2014; *N* = 253,280), both of which did not include UKB.

### LD Score Regression

SNP *h^2^* was estimated using the version of LDSC (Bulik-Sullivan et al., 2015) implemented in the Genomic SEM R package (Grotzinger et al., 2019). Because, the majority of our simulations and empirical data analyses capitalized on genotypes from individuals of European-like ancestry from UKB, we used European-like ancestry LD scores from the Pan-UK Biobank reference panel (Karczewski et al., 2025). We selected this reference panel following preliminary simulation analyses using UKB genotypes in which application of LDSC using the European-like ancestry 1000 Genomes reference panel (1000 Genomes Project Consortium, 2015) produced modest overestimates of SNP *h^2^*. Specifically, across 15 simulation replications with a SNP *h^2^_true_* of 0.20, the mean LDSC estimate was 0.219 using the 1000 Genomes reference panel, compared with 0.197 using Pan-UK Biobank. We therefore used Pan-UK Biobank LD scores for subsequent analyses.

### PGI Construction Methods

Below we describe each PGI method employed and the associated LD reference panel, when relevant. To ensure comparability across PGI construction methods, we constructed PGIs from the 749,619 SNPs that were common across all LD reference panels and available in UKB after merging.

#### Unadjusted

PGI Unadjusted was constructed by weighting each allele by its corresponding marginal GWAS effect estimate, without LD modeling, clumping, or shrinkage.

#### PRS-CS-auto

PRS-CS-auto constructs PGIs using a Bayesian regression framework that applies continuous shrinkage priors to SNP effect sizes while explicitly accounting for LD among variants. The method uses GWAS summary statistics together with an external LD reference panel to estimate posterior SNP effect sizes under a global-local shrinkage prior, allowing effect sizes to be adaptively shrunk according to the strength of evidence for association. We used the UKB European reference panel provided with PRS-CS, which is based on 1,120,697 HapMap3 variants and captures LD patterns across the genome. In the auto implementation, the global shrinkage parameter controlling the overall sparsity of the genetic architecture is estimated directly from the data, eliminating the need for an external validation sample or parameter tuning. Posterior SNP effect estimates were obtained using Markov chain Monte Carlo sampling, and PGIs were constructed using the posterior mean effect sizes. This approach allows the degree of shrinkage applied to SNP effects to be learned from the observed GWAS summary statistics while accounting for LD among nearby variants.

#### SBayesR

SBayesR performs Bayesian multiple regression using GWAS summary statistics and an external LD reference panel, modeling SNP effect sizes under a finite mixture prior consisting of a point mass at zero and multiple normal distributions with increasing variances. We specified mixture proportions of 0.95, 0.02, 0.02, and 0.01 across four components, with corresponding variance scaling parameters of 0, 0.01, 0.1, and 1, such that the majority of SNPs were assumed to have null effects while allowing a small proportion to have progressively larger non-zero effects. Posterior SNP effect sizes were estimated via Markov chain Monte Carlo sampling, and PGIs were constructed using the posterior mean effect estimates. LD was modeled using the UK Biobank 10K HapMap3 shrunk sparse LD reference panel based on 1,094,841 variants.

#### Clumping

PGI weights were constructed using LD-based clumping. Clumping did not use a separate precomputed LD reference panel; LD was estimated directly from the genotype data used in the clumping procedure via PLINK2. For each discovery half, GWAS summary statistics were subjected to LD clumping, in which SNPs were ranked by statistical significance and the most significant variant within a given genomic window was retained while nearby variants in LD above a specified *r^2^* threshold were removed. No p-value threshold was imposed at the clumping stage, such that all SNPs were eligible for selection and retention was determined by LD structure rather than significance filtering.

We used data on educational attainment, height, and verbal-numeric reasoning within the UKB dataset to evaluate the sensitivity of results to clumping parameters. We examined multiple combinations of physical window sizes (1 Mb, 3 Mb, and 5 Mb) and LD thresholds (*r^2^ =* 0.1, 0.05, 0.01; see **Supplementary Table S4**). Across parameter combinations, the highest error-corrected prediction accuracies for educational attainment and height were obtained from PGIs constructed using a 5 Mb window and an LD threshold of *r²* = 0.01. Although this specification produced the lowest error-corrected prediction accuracy for verbal-numeric reasoning, estimates were very similar across parameter combinations. We therefore retained the 5 Mb window and LD threshold of *r²* = 0.01 for all clumping-based PGIs reported throughout the main text, including those constructed for other phenotypes and simulated phenotypes. Results from the alternative parameter specifications are reported in **Supplementary Table S4**.

#### LDpred2-auto and LDPred2-inf

For LDpred2, we used LD estimates for 1,054,330 HapMap3 variants derived from 362,320 individuals of European-like ancestry in the UKB. Whereas LDpred2-inf assumes an infinitesimal genetic architecture in which all variants contribute to the phenotype, LDpred2-auto uses a Bayesian spike-and-slab model that allows some variants to have zero effects. Specifically, LDpred2-auto estimates the proportion of causal variants and SNP heritability directly from the GWAS summary statistics using a Gibbs sampling procedure, rather than requiring these parameters to be specified or tuned using an external validation sample (Privé et al., 2020). PGIs were constructed using the resulting posterior mean SNP effect estimates.

## Data Availability

All data produced in the present study are available upon reasonable request.

https://utexas.app.box.com/s/ydkfr84rjr5w60xd3zjjlcsc54o4ls4q

## Acknowledgements

E.M.T.-D is a member of the Population Research Center (PRC) and the Center on Aging and Population Sciences (CAPS) at The University of Texas at Austin, which are supported by NIH grants P2CHD042849 and P30AG066614, respectively. M.G.N. is a member of the MRC Integrative Epidemiology Unit at the University of Bristol, which is supported by the Medical Research Council and the University of Bristol grants MC_UU_00032/1 and MC_UU_00032/7, respectively. Analyses of data from the UK Biobank (UKB) were performed under approved project 10279. The Health and Retirement Study (HRS) is sponsored by the National Institute on Aging (grant U01AG009740) and is conducted by the University of Michigan. We are grateful to Aysu Okbay for providing us with UKB-left-out summary statistics for educational attainment.

## Data and Code Availability

Summary statistics from noise GWAS conducted in the UKB and the HRS, which can be used to create synthetic splits of GWAS summary statistics, are available at https://utexas.box.com/s/ydkfr84rjr5w60xd3zjjlcsc54o4ls4q. An R script demonstrating the synthetic splitting procedure is also available at the same link.

## Author contributions

Y.D., R.E.H., M.G.N. and E.M.T.-D. conceived the study. Y.D., M.G.N. and E.M.T.-D. designed the study and drafted and revised the manuscript. Y.D., R.E.H. and E.M.T.-D. conducted simulation analyses. Y.D. and T.S. conducted analyses of empirical data. G.W.E. and D.L.-C. provided input on the analysis plan and contributed to the interpretation of the results. M.G.N. and E.M.T.-D supervised this research. All authors approved the final version of the manuscript.

---

**Extended Data Figure 1.**
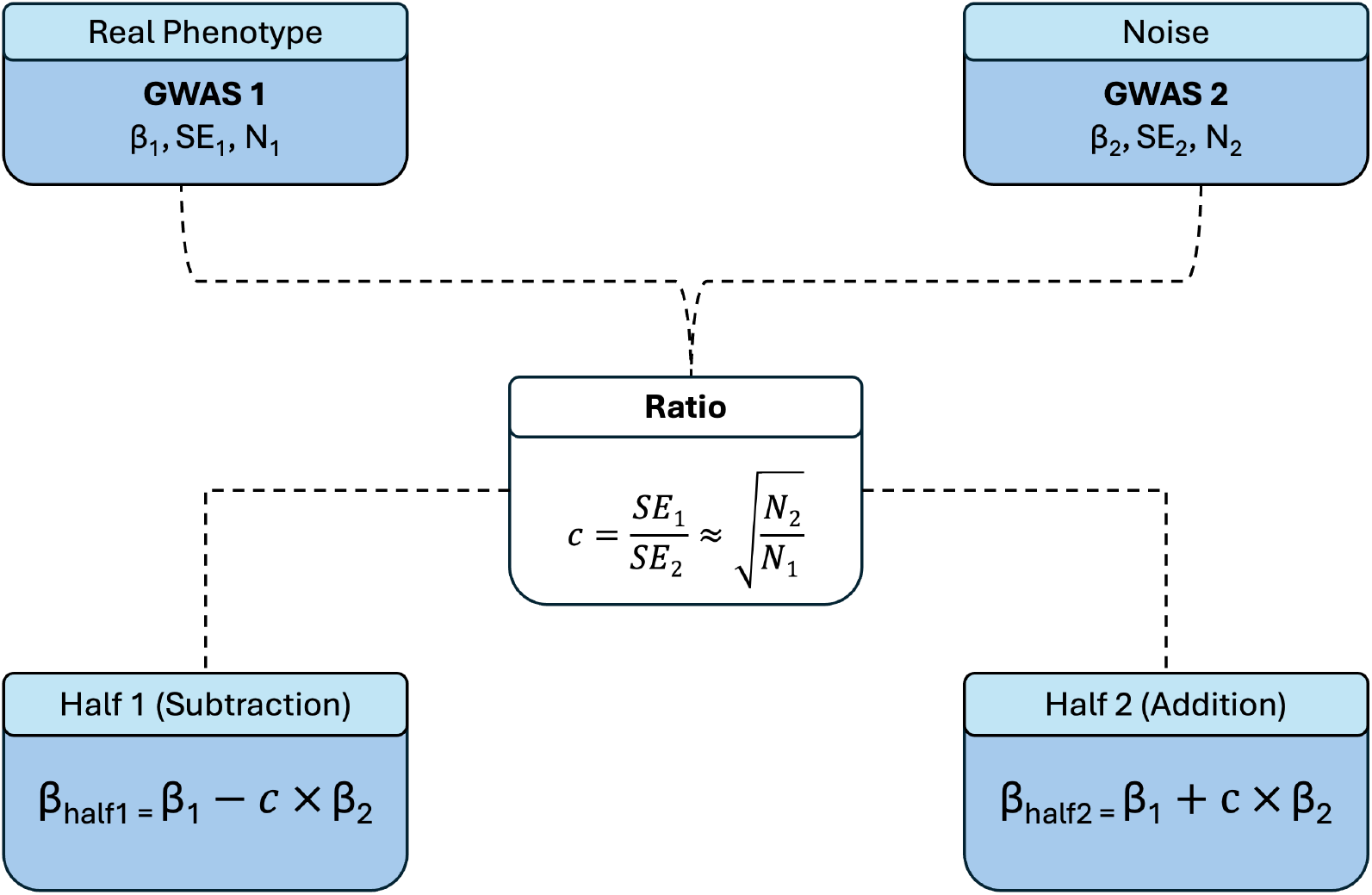
Diagram of pipeline for synthetically splitting a single set of GWAS summary statistics. GWAS summary statistics for a measured phenotype of interest (GWAS 1) are split using GWAS summary statistics formed by performing GWAS on a randomly generated pseudo phenotype i.e., noise phenotype (GWAS 2). For each SNP, the estimate from the noise GWAS (β₂) is rescaled using the ratio of the standard errors from the two GWASs (c = SE₁/SE₂), which is approximately equal to the square root of the ratio of their sample sizes. The rescaled noise effect is then subtracted from or added to the estimate (β₁) for the corresponding SNP from the GWAS of the measured phenotype. This produces two approximately independent sets of GWAS summary statistics, each with an associated sample size of half the original GWAS (N₁/2) and standard errors inflated by a factor of √2. The resulting synthetic halves can then be used to construct independent PGIs without requiring access to individual-level data from the original GWAS.

## References

1000 Genomes Project Consortium. (2015). A global reference for human genetic variation. Nature, 526(7571), 68.

Becker, J., Burik, C. A., Goldman, G., Wang, N., Jayashankar, H., Bennett, M., … & Okbay, A. (2021). Resource profile and user guide of the Polygenic Index Repository. Nature Human Behaviour, 5(12), 1744–1758.

Bulik-Sullivan, B. K., Loh, P. R., Finucane, H. K., Ripke, S., Yang, J., Patterson, N., … & Neale, B. M. (2015). LD Score regression distinguishes confounding from polygenicity in genome-wide association studies. Nature Genetics, 47(3), 291–295.

Bycroft, C., Freeman, C., Petkova, D., Band, G., Elliott, L. T., Sharp, K., … & Marchini, J. (2018). The UK Biobank resource with deep phenotyping and genomic data. Nature, 562(7726), 203–209. 10.1038/s41586-018-0579-z

Chang, C. C., Chow, C. C., Tellier, L. C., Vattikuti, S., Purcell, S. M., & Lee, J. J. (2015). Second-generation PLINK: rising to the challenge of larger and richer datasets. Gigascience, 4(1), s13742–015.

Choi, S. W., & O’Reilly, P. F. (2019). PRSice-2: Polygenic Risk Score software for biobank-scale data. Gigascience, 8(7), giz082.

Choi, S. W., Mak, T. S. H., & O’Reilly, P. F. (2020). Tutorial: a guide to performing polygenic risk score analyses. Nature Protocols, 15(9), 2759–2772.

Demange, P. A., Malanchini, M., Mallard, T. T., Biroli, P., Cox, S. R., Grotzinger, A. D., … & Nivard, M. G. (2021). Investigating the genetic architecture of noncognitive skills using GWAS-by-subtraction. Nature Genetics, 53(1), 35–44.

DiPrete, T. A., Burik, C. A., & Koellinger, P. D. (2018). Genetic instrumental variable regression: Explaining socioeconomic and health outcomes in nonexperimental data. Proceedings of the National Academy of Sciences, 115(22), E4970–E4979.

Frach, L., Rijsdijk, F., Hannigan, L. J., Dudbridge, F., & Pingault, J. B. (2026). Genetic sensitivity analysis: estimating genetic confounding and environmentally mediated genetic effects using multiple exposures. medRxiv, 2026–07.

Furrer, R. A., & Turkheimer, E. N. (2026). Polygenic risk scores are not genetic predispositions. Nature Human Behaviour, 1–2.

Gazal, S., Finucane, H. K., Furlotte, N. A., Loh, P. R., Palamara, P. F., Liu, X., … & Price, A. L. (2017). Linkage disequilibrium–dependent architecture of human complex traits shows action of negative selection. Nature Genetics, 49(10), 1421–1427.

Ge, T., Chen, C. Y., Ni, Y., Feng, Y. C. A., & Smoller, J. W. (2019). Polygenic prediction via Bayesian regression and continuous shrinkage priors. Nature Communications, 10(1), 1776.

Grotzinger, A. D., Rhemtulla, M., de Vlaming, R., Ritchie, S. J., Mallard, T. T., Hill, W. D., … & Tucker-Drob, E. M. (2019). Genomic structural equation modelling provides insights into the multivariate genetic architecture of complex traits. Nature Human Behaviour, 3(5), 513–525.

Hoerl, A. E., & Kennard, R. W. (1970). Ridge regression: Biased estimation for nonorthogonal problems. Technometrics, 12(1), 55–67.

Karczewski, K. J., Gupta, R., Kanai, M., Lu, W., Tsuo, K., Wang, Y., … & Martin, A. R. (2025). Pan-UK Biobank genome-wide association analyses enhance discovery and resolution of ancestry-enriched effects. Nature Genetics, 57(10), 2408–2417.

Lloyd-Jones, L. R., Zeng, J., Sidorenko, J., Yengo, L., Moser, G., Kemper, K. E., … & Visscher, P. M. (2019). Improved polygenic prediction by Bayesian multiple regression on summary statistics. Nature Communications, 10(1), 5086.

Malawsky, D. S., Hegemann, L., Wootton, O., Havdahl, A. K., Sunde, H. F., & Martin, H. C. (2026). Phantom genetic nurture: assortative mating accounts for most of the apparent association between parental genotypes and childhood cognitive performance. bioRxiv, 2026–09.

Mbatchou, J., Barnard, L., Backman, J., Marcketta, A., Kosmicki, J. A., Ziyatdinov, A., … & Marchini, J. (2021). Computationally efficient whole-genome regression for quantitative and binary traits. Nature Genetics, 53(7), 1097–1103.

Okbay, A., Wu, Y., Wang, N., Jayashankar, H., Bennett, M., Nehzati, S. M., … & Esko, T. (2022). Polygenic prediction of educational attainment within and between families from genome-wide association analyses in 3 million individuals. Nature Genetics, 54(4), 437–449.

Pingault, J. B., Rijsdijk, F., Schoeler, T., Choi, S. W., Selzam, S., Krapohl, E., … & Dudbridge, F. (2021). Genetic sensitivity analysis: Adjusting for genetic confounding in epidemiological associations. PLoS Genetics, 17(6), e1009590.

Privé, F., Arbel, J., & Vilhjálmsson, B. J. (2020). LDpred2: better, faster, stronger. Bioinformatics, 36(22-23), 5424–5431.

Rosseel, Y. (2012). lavaan: An R package for structural equation modeling. Journal of statistical software, 48(1), 1–36.

Schwaba, T., Clapp Sullivan, M. L., Akingbuwa, W. A., Ilves, K., Tanksley, P. T., Williams, C. M., … & Tucker-Drob, E. M. (2026). Robust inference and correlates from genetic associations with personality. Nature, 1–11.

Sinnott-Armstrong, N., Naqvi, S., Rivas, M., & Pritchard, J. K. (2021). GWAS of three molecular traits highlights core genes and pathways alongside a highly polygenic background. Elife, 10, e58615.

Spearman, C. (1904). The proof and measurement of association between two things. The American Journal of Psychology, 15(1), 72–101.

Tibshirani, R. (1996). Regression shrinkage and selection via the lasso. Journal of the Royal Statistical Society: Series B, 58(1), 267–288.

Tucker-Drob, E. M. (2017). Measurement error correction of genome-wide polygenic scores in prediction samples. BioRxiv, 165472.

Wang, J. Y., Lin, N., Zietz, M., Mares, J., Smith, O. S., Rathouz, P. J., & Harpak, A. (2026). Three open questions in polygenic score portability. Nature Communications, 17(1), 942.

Wang, X., Walker, A., Revez, J. A., Ni, G., Adams, M. J., McIntosh, A. M., … & Arolt, V. (2023). Polygenic risk prediction: why and when out-of-sample prediction R2 can exceed SNP-based heritability. The American Journal of Human Genetics, 110(7), 1207–1215.

Wood, A. R., Esko, T., Yang, J., Vedantam, S., Pers, T. H., Gustafsson, S., … & Kratzer, W. (2014). Defining the role of common variation in the genomic and biological architecture of adult human height. Nature Genetics, 46(11), 1173–1186.

Wray, N. R., Goddard, M. E., & Visscher, P. M. (2007). Prediction of individual genetic risk to disease from genome-wide association studies. Genome research, 17(10), 1520.

Wray, N. R., Yang, J., Hayes, B. J., Price, A. L., Goddard, M. E., & Visscher, P. M. (2013). Pitfalls of predicting complex traits from SNPs. Nature Reviews Genetics, 14(7), 507–515.

Yang, J., Lee, S. H., Goddard, M. E., & Visscher, P. M. (2011). GCTA: a tool for genome-wide complex trait analysis. The American Journal of Human Genetics, 88(1), 76–82.

Zeng, J., De Vlaming, R., Wu, Y., Robinson, M. R., Lloyd-Jones, L. R., Yengo, L., … & Yang, J. (2018). Signatures of negative selection in the genetic architecture of human complex traits. Nature Genetics, 50(5), 746–753.

Zheng, Z., Liu, S., Sidorenko, J., Wang, Y., Lin, T., Yengo, L., … & Zeng, J. (2024). Leveraging functional genomic annotations and genome coverage to improve polygenic prediction of complex traits within and between ancestries. Nature Genetics, 56(5), 767–777.

